# Standardization and Age-Distribution of COVID-19: Implications for Variability in Case Fatality and Outbreak Identification

**DOI:** 10.1101/2020.04.09.20059832

**Authors:** David N. Fisman, Amy L. Greer, Ashleigh R. Tuite

## Abstract

**Background:** Epidemiological data from the COVID-19 pandemic has demonstrated variability in attack rates by age, and country-to-country variability in case fatality ratio (CFR).

**Objective:** To use direct and indirect standardization for insights into the impact of age-specific under-reporting on between-country variability in CFR, and apparent size of COVID-19 epidemics.

**Design:** Post-hoc secondary data analysis (“case studies”), and mathematical modeling.

**Setting:** China, global.

**Interventions:** None.

**Measurements:** Data were extracted from a sentinel epidemiological study by the Chinese Center for Disease Control (CCDC) that describes attack rates and CFR for COVID-19 in China prior to February 12, 2020. Standardized morbidity ratios (SMR) were used to impute missing cases and adjust CFR. Age-specific attack rates and CFR were applied to different countries with differing age structures (Italy, Japan, Indonesia, and Egypt), in order to generate estimates for CFR, apparent epidemic size, and time to outbreak recognition for identical age-specific attack rates.

**Results:** SMR demonstrated that 50-70% of cases were likely missed during the Chinese epidemic. Adjustment for under-recognition of younger cases decreased CFR from 2.4% to 0.8% (assuming 50% case ascertainment in older individuals). Standardizing the Chinese epidemic to countries with older populations (Italy, and Japan) resulted in larger apparent epidemic sizes, higher CFR and earlier outbreak recognition. The opposite effect was demonstrated for countries with younger populations (Indonesia, and Egypt).

**Limitations:** Secondary data analysis based on a single country at an early stage of the COVID-19 pandemic, with no attempt to incorporate second order effects (ICU saturation) on CFR.

**Conclusion:** Direct and indirect standardization are simple tools that provide key insights into between-country variation in the apparent size and severity of COVID-19 epidemics.

**Funding:** The research was supported by a grant to DNF from the Canadian Institutes for Health Research (2019 COVID-19 rapid researching funding OV4-170360).

## Introduction

Knowledge and understanding related to COVID-19 are evolving rapidly, thanks in no small part to outstanding epidemiological work done under challenging conditions in recent months (1). A report on 44,672 confirmed COVID-19 cases from mainland China helped delineate early understanding of the outbreak’s epidemiology. More recent mathematical models help fill in some of the informational gaps, by inferring the underlying processes, including the occurrence of “silent”, unrecognized infections, that must have driven this epidemic (2). Modeling is an important tool for understanding epidemic processes, but disease modeling expertise is not universally available. A much more basic epidemiological tool (standardization) (3, 4) can be used to provide important insights into both seen and unseen aspects of epidemics, and to project the likely characteristics and impacts of the same epidemic process, if it were to unfold in other populations.

We were struck by the absence of reported COVID-19 cases in younger individuals in early reports from China. A pandemic disease is defined by the novelty of the pathogen and absence of population-level immunity, such that all age groups in a population should be equally susceptible to infection. Inasmuch as more severe cases are more likely to be recognized, the under-recognition of disease in younger individuals serves as a metric for differential disease severity by age, and also provides important information that can be used to adjust case fatality ratios for likely under-reporting. Furthermore, simple approaches to quantify under-reporting can inform public health prevention strategies, because if unrecognized cases are extremely common, control methods that focus on identification of cases, isolation and quarantine alone are likely to fail.

We sought to use simple epidemiological tools, such as direct and indirect standardization (i.e., calculation of standardized morbidity ratios) to gain insights into likely disease under-reporting and case fatality in mainland China. We then applied these insights to infer likely differences in disease severity (based on CFR), and detection of epidemics occurring in countries *outside* mainland China.

## Methods

### Data Sources

COVID-19 case counts by age were based on confirmed cases, by age, reported in (5). 2020 country population projections for China by age were obtained from the United Nations using the UNWPP package in R (6, 7). While the Chinese COVID-19 epidemic was centered on the province of Hubei, the epidemic rapidly spread to involve all Chinese provinces. Therefore, we used the total Chinese population data by age to calculate age-specific cumulative incidence over the initial 9 weeks of the epidemic. We used these initial observations to perform all subsequent analyses.

### Standardized Morbidity Ratios

We calculated overall cumulative incidence per 100,000 population in the 66- days from December 8, 2019 (the date of onset in the first recognized human COVID-19 case) to February 11, 2020 (8). Crude and age-specific cumulative incidence were calculated as the ratio of case numbers to population size. Standardized morbidity ratios (SMR) were then calculated as 100 × (observed cases/expected cases) where expected cases are the product of crude cumulative incidence and the population size of a given age group (4).

### Under-Ascertainment of Younger Cases and Implications for Case-Fatality

Given that COVID-19 is an emerging communicable disease and there is no pre-existing immunity in the population, attack rates should be similar across age groups, or possibly even higher in children due to their more intense contact structure (9). The elevated SMR in older age groups, combined with their higher case fatality, is suggestive of increased case ascertainment in this group due to greater clinical severity. Indeed, when active case finding has been performed for pediatric cases, attack rates in younger groups have been similar to those in the older age groups. We examined a series of “case studies” where incidence in older individuals (age > 59) was assumed to be measured accurately, and cumulative incidence in older individuals was then applied to younger age groups to generate estimates of the fraction of cases under-ascertained in these age groups. We then revised the expected case fatality proportions based on case counts adjusted for likely under-reporting in younger individuals.

### Population Standardization, Case Fatality and Observed Outbreak Size

We evaluated the anticipated size, timing, and impact of an epidemic with identical age-specific cumulative incidence and case fatality as observed in China but applied to four countries outside of China. We standardized to countries and areas with older age than China (Japan, Italy) and younger age (Indonesia, Egypt) as a means of isolating the impact of age structure on outbreak characteristics. While somewhat arbitrary, these regions have all either been impacted by COVID-19 to some degree (Japan, and Italy) (10-12); have had large numbers of exported cases without large national epidemics (Egypt)(13); or have been notable for the relatively limited number of cases identified notwithstanding close links to China (Indonesia) (14).

Since China’s large population size results in a far larger epidemic for a given incidence, we used a ratio-of-ratio approach. The ratio of population in the other, comparator country (P_O_) to the Chinese population (P_C_) was defined as R_P_ = P_O_/P_C_. The ratio of the observed epidemic size in the other, comparator country (E_0_) to observed Chinese epidemic size (E_C_) was defined as R_E_ = E_O_/E_C_. The ratio of ratios was thus R_E_/R_P_, and can be interpreted as the relative apparent outbreak size when an outbreak with identical age-specific attack rates occurs in a population with an age-structure that differs from that of China.

### Age Structure and Outbreak Detection

We estimated the incidence of observed infection among susceptible older individuals (age > 59) in the Chinese population required for the observed epidemic to have taken place over 66 days using the relation λ = −ln(1-P)/t. This hazard was then applied to 1) the Chinese population, and 2) the populations of the other four “case study” countries, over a 66-day period under the assumption that the most severe illness would be seen in those aged > 59 years. We modeled time to observation of deaths by modeling time to symptoms, severe pneumonia, ICU admission, and death using parameter estimates presented in **Table 1**, assuming exponential failure time.

**Table 1.** Parameters Used for Time-To-Death Estimates.

| Parameter |  | Estimate | Reference |
| --- | --- | --- | --- |
| Proportions |  |  |  |
|  | Severe pneumonia | 0.15 | (23) |
|  | ICU requirement with severe pneumonia | 0.20 | (23) |
|  | Death in ICU | 0.62 | (24) |
| Average duration (days) |  |  |  |
|  | Incubation period | 6 | (25) |
|  | Time from onset to hospitalization | 7 | (26) |
|  | Time from hospitalization to ICU | 3 | (26) |
|  | Time from ICU admission to death | 25 | (25) |
| Force of infection ($\lambda$) | | $8.44 \times 10^{-7}$ | Calculated based on (5). |

**NOTE:** ICU, intensive care unit.

## Results

Based on data in (8), the crude cumulative incidence of observed COVID-19 in mainland China up until February 11, 2020 was 3.1 per 100,000. By contrast, cumulative incidence in those aged > 59 years was 5.6 per 100,000. Age-specific cumulative incidence and SMR by age are presented in **Table 2** and **Supplementary Figure 1**. It can be seen that SMR for age groups < 50 was substantially lower than that in older age groups and most deaths were also observed in older age groups (**Table 2**). When we assumed complete or near complete ascertainment of cases in individuals aged >59, and adjusted incidence in younger age groups accordingly, the adjusted CFR fell, and was 0.8% if we assume that only 50% of older cases were ascertained (**Figure 1**). Even if all cases were ascertained in older individuals, it was estimated that 46% of total cases were missed; if only 50% of older cases were ascertained it was estimated that 75% of cases were missed (**Figure 1**).

**Table 2:** Epidemiological Characteristics of China’s COVID-19 Epidemic to February 11, 2020.

| **Age**<br>**Group** | **Cases** | **Deaths** | **Case**<br>**Fatality** | **Population**<br>**(millions)** | **Cumulative Incidence\*** | **SMR** |
| --- | --- | --- | --- | --- | --- | --- |
| 0-9 | 416 | 0 | 0 | 170.7 | 0.24 | 7.9 |
| 10-19 | 549 | 1 | 0.002 | 166.6 | 0.33 | 10.6 |
| 20-29 | 3619 | 7 | 0.002 | 185.1 | 1.95 | 63.0 |
| 30-39 | 7600 | 18 | 0.002 | 228.8 | 3.32 | 107.0 |
| 40-49 | 8571 | 38 | 0.004 | 216.1 | 3.97 | 127.8 |
| 50-59 | 10008 | 130 | 0.013 | 222.2 | 4.50 | 145.1 |
| 60-69 | 8583 | 309 | 0.036 | 151.7 | 5.66 | 182.3 |
| 70-79 | 3918 | 312 | 0.080 | 71.5 | 5.48 | 176.6 |
| 80+ | 1408 | 208 | 0.148 | 26.6 | 5.29 | 170.4 |

**NOTE:** SMR, standardized morbidity ratio.
\*per 100,000 population.

**Figure 1.**
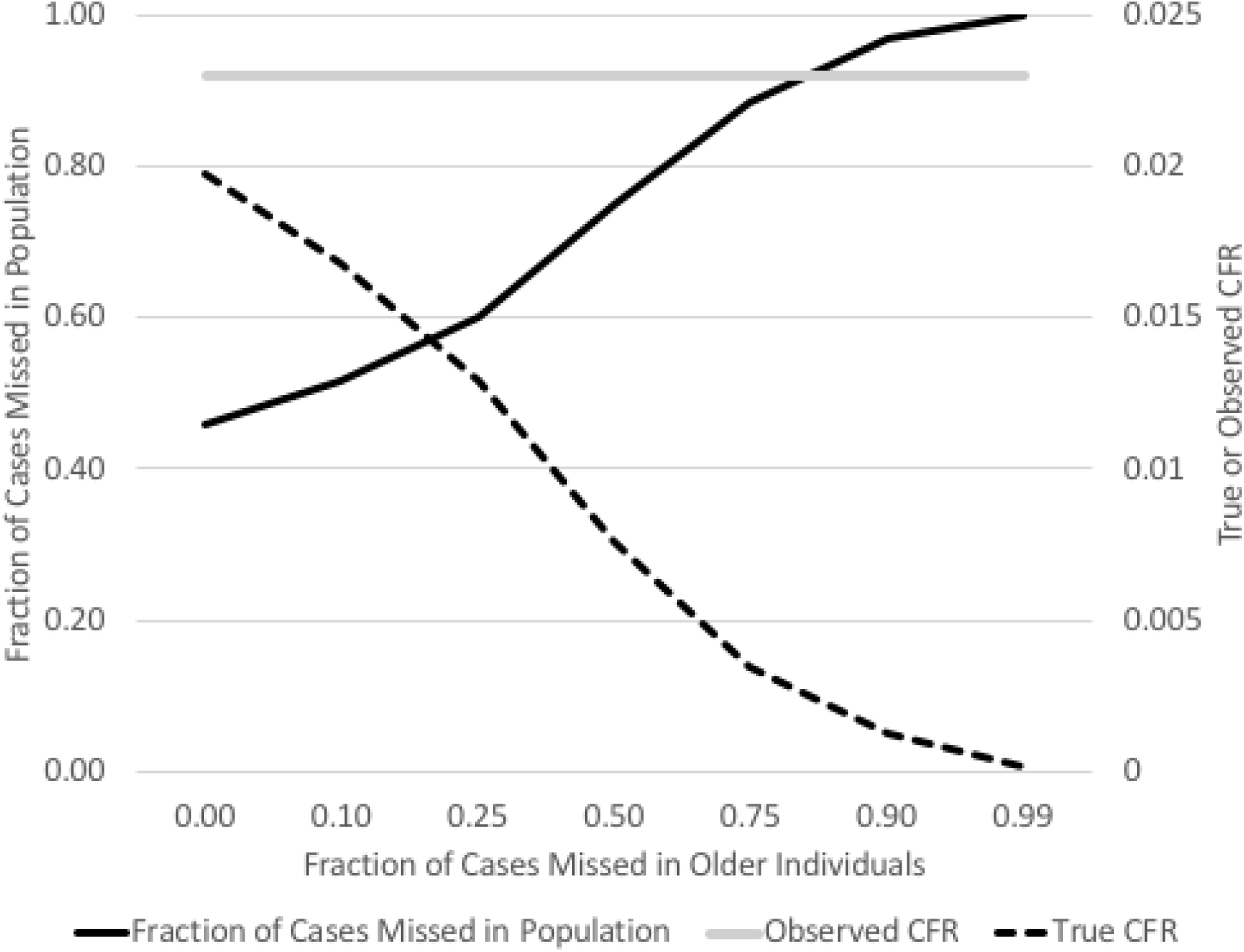
Case Fatality and Fraction of Cases Missed Under Varying Assumptions of Reporting Completeness in Older Individuals. Estimates of the fraction of cases missed in the population as a whole (black solid curve), and true case-fatality ratio (CFR) (black dashed curve), as a function of the fraction of cases missed in older adults who are assumed to be ascertained with the greatest accuracy. Decreasing case ascertainment in older adults implies an even higher fraction of cases are missed in the population as a whole, and CFR is lower than observed.

When the Chinese epidemic was age-standardized using population pyramids from other countries, standardization to younger populations (Indonesia, Egypt and Iran) markedly reduced CFR, while adjustment to older countries or regions (Japan, Italy) elevated CFR (**Table 3**). The ratio-of-ratios, R_E_/R_P_, was less than 1 for countries with younger populations, but greater than 1 for countries with older populations. In other words, apparent epidemics, adjusted for population size, would be expected to be smaller in countries with younger populations (shorter life expectancy) than in those with older populations (increased life expectancy), even with identical age-specific attack rates.

**Table 3:** Direct Standardization of Mainland China’s COVID-19 Epidemic to Other Countries and Regions.

| Country/Region | Country-Standardized<br>CFR (%) | Epidemic<br>Size Ratio<br>(R<sub>E</sub>)* | Population<br>Size Ratio<br>(R<sub>P</sub>)* | R<sub>E</sub>/R<sub>P</sub> |
| --- | --- | --- | --- | --- |
| Mainland China | 2.3 | --- | --- | --- |
| Egypt | 1.6 | 0.05 | 0.07 | 0.69 |
| Indonesia | 1.7 | 0.15 | 0.19 | 0.81 |
| Italy | 3.9 | 0.05 | 0.04 | 1.15 |
| Japan | 4.4 | 0.10 | 0.09 | 1.18 |

**NOTE:** CFR, case fatality ratio.
\*Compared to Mainland China.

When we simulated the mainland China epidemic in other countries, we found that at any threshold of deaths required for outbreak detection, outbreaks would be detected more quickly in countries with high life expectancy, and more slowly in those with low life expectancy (**Figure 2** and **Online Appendix** (https://art-bd.shinyapps.io/time_to_outbreak_detection/).

**Figure 2.**
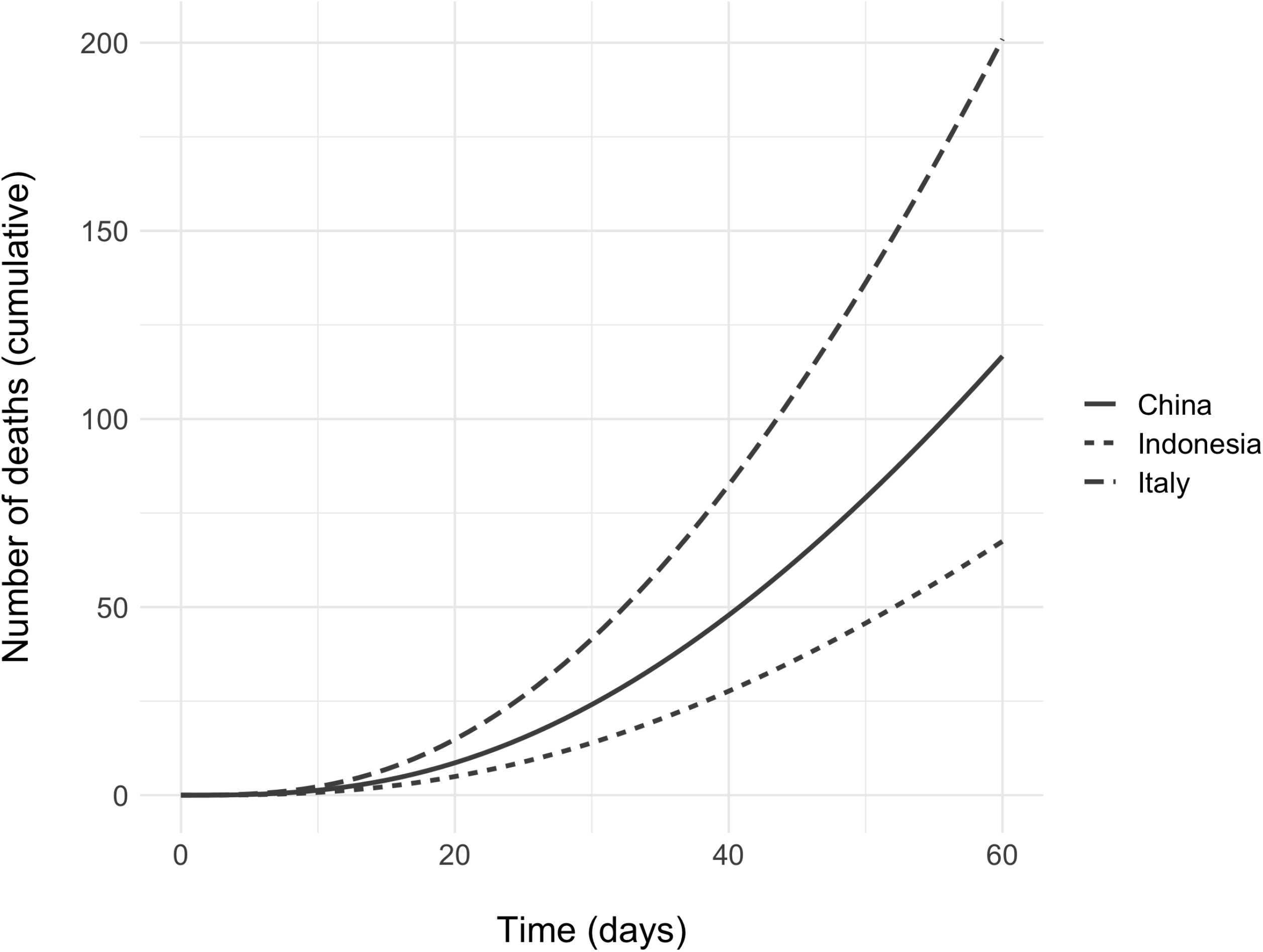
Model Describing Differential Time To Recognition of COVID-19 Outbreaks in Countries with Different Age Structures. Outbreaks with identical age-specific attack rates, and otherwise identical characteristics, were simulated in countries with intermediate (China), old (Italy) and young (Indonesia) populations. It can be seen that for any threshold of deaths that must be exceeded for an outbreak to be recognized, older countries will be identified before younger countries. Model details are as described in the text.

## Discussion

As the COVID-19 pandemic has expanded its reach, the role of unrecognized infection has received increased scrutiny (2, 15). While individuals with unrecognized infection may be important in the epidemic’s spread, those with more severe illness are more likely to be recognized clinically, and more likely to be referred for virological testing, a practice which the age distribution of identified cases in China early in the pandemic, foretold (8).

Age-related increases in severity, which may be confounded by increasing prevalence of chronic medical conditions with age, are now well described in countries outside China (16, 17). Greater recognition of individuals with more severe illness, and undercounting of those with mild infection, is likely to inflate apparent case fatality. While serological testing will ultimately help determine the true infection fatality ratio for COVID-19, estimates of undercounting may be derived if it is assumed that all in the population, regardless of age, are equally vulnerable to infection. We demonstrate such an approach in this paper.

The key driver of pandemic disease is a fully susceptible population; novel pathogens have higher reproduction numbers when they first emerge but the number drops once some proportion of the population has become immune (18). This leads to very high attack rates early in a pandemic. Furthermore, vulnerability to infection should be equally distributed across the population, with incidence expected to be highest in children, who have the highest rates and intensity of person-to-person contact. As such, an absence of pediatric cases in national reporting data represents an index of under-reporting rather than immunity to infection and can be used as a means of quickly adjusting models for under-reported fractions through simple, easily applied methods such as direct and indirect standardization, which we employ here. Bayesian methods provide a more computationally intensive and more technically challenging approach to the same problem (2).

The extraordinary case-fatality in the COVID-19 pandemic (as high as 10-12% in Spain and Italy as of April 3, 2020) (19), also underscores the unusual epidemiology of pandemics, since with endemic diseases (and some pandemics, such as the 2009 (H1N1) influenza A pandemic) early life immune experience protects those who would be vulnerable to severe disease conditional on infection (i.e., older individuals), while permitting infection of younger individuals less likely to experience severe disease (20). While case-fatality is driven at least in part by the extent of testing, standardizing these epidemics to different populations (in effect, letting an identical epidemic run out in a different population) allows us to see that demographic structure alone can explain many between country differences in apparent epidemic size and case fatality. Adjusting for population size, identical epidemics will appear larger and more severe in “older” countries (like those in Western Europe) and smaller and milder in “younger” countries (like Egypt, and Indonesia).

A key limitation of this work is that much of the work focusses on an epidemic in a single country, at an early point in the COVID-19 pandemic. Indeed, the observable case-fatality in China now approximates 4%, rather than 2.4% as reported earlier, which is likely to reflect lags between clinical onset and death from COVID-19, especially in individuals who receive intensive care with mechanical ventilation. We have, furthermore, not attempted to incorporate second order effects, such as the resulting rapid saturation of ICU resources, with resultant upwards inflection in case fatality, in countries with older populations (e.g. Italy). Such effects may be operative in the devastating COVID- 19 epidemics in Western Europe, which have CFR well beyond what our standardization of the Chinese epidemic data would predict.

In conclusion, we find that standardization, both direct and indirect, provides a simple, widely understood toolbox for explaining and understanding several of the unusual features of COVID-19, including under-representation of pediatric cases and geographic variability in apparent epidemic size and severity (measured as CFR). While we are living in frightening and emotionally charged times, we suggest that demographic variation, rather than misrepresentation (21, 22), is likely to explain much of the between-country variability seen in the current pandemic.

## Data Availability

All data used in the paper are publicly available.

## Figure Legends

**Supplementary Figure 1. Observed Cumulative Incidence, Deaths and Standardized Morbidity Ratios for Mainland China COVID-19 Epidemic**. Figure is a graphical representation of data presented in **Table 2**. SMR are estimated as 100 × observed incidence divided by expected incidence, which in the context of a pandemic is approximately equal in all age groups, or somewhat higher in younger individuals.

## Notes

**Competing interests:** none

### Competing Interest Statement

The authors have declared no competing interest.

